# Evaluation of model–informed precision dosing of cefepime in critically ill patients: a French before– after study

**DOI:** 10.1101/2025.08.28.25334648

**Authors:** Giuseppe Balice, Soizic Percevault, Sabine Cohen, Romain Garreau, Florent Wallet, Arnaud Friggeri, Sylvain Goutelle

## Abstract

Cefepime is widely used in the intensive care unit (ICU) for the treatment of complicated Gram-negative infections. Cefepime is a candidate for therapeutic drug monitoring (TDM) and model-informed precision dosing (MIPD), especially in critically ill patients, notably because of its concentration-dependent neurological toxicity. In this study, we aimed to evaluate the impact of the implementation of cefepime MIPD on PK/PD targets attainment in intensive care. We performed a monocentric, retrospective, before‒after study, including all adult patients hospitalized in our ICU and for whom at least two cefepime plasma through concentrations (C_min_) were measured on separate days. The main endpoint was a C_min_ between 4×MIC (or 10mg/L if the MIC was unavailable) and 20mg/L. We modelled the odds of target attainment via a mixed-effect logistic model and the rate of target attainment with a spline of time.

A total of 281 patients were included, of whom 121 were in the MIPD group and 160 in the control group. Median age, weight and follow-up duration were 65 years, 76 kg and 3 days respectively. The two most common infections were pneumonia (n=239) and peritonitis (n=16). A total of 728 cefepime through concentrations were collected. MIPD was non-significantly associated with higher odds of being in the therapeutic range (aOR 1.40 [0.88 – 2.22]) and significantly associated with lower odds of being over-exposed (aOR 0.58 [0.35 – 0.96]). HR of target attainment was 1.1 [0.7 – 1.9] at day 1 and 1.6 [0.9 – 2.9] at day 7.

In summary, we demonstrated that cefepime MIPD in the ICU reduces the risk of over-exposure, and may be beneficial on the odds and on the rate of PK/PD targets attainment.

**What is already known about this subject:**

- Cefepime is a candidate for TDM/MIPD programs, with established PK/PD targets for both antimicrobial efficacy and neurological toxicity (100% *f* T >4×MIC and C_min_>20mg/L, respectively).
- Previous randomized controlled trials (RCT) of beta-lactams model-informed precision dosing (MIPD) versus standard dosing approaches failed to demonstrate a superiority of MIPD in clinical endpoints attainment, and provided conflicting results on pharmacokinetic target attainment.

**What this study adds:**

- Cefepime MIPD in the intensive care units reduces the odds of being over-exposed, and thus, possibly, the risk of elicit dose-dependent neurological toxicity.
- Further RCT evaluating MIPD efficacy should focus on evaluating target attainment rate instead of probability alone.

## Introduction

Cefepime is a fourth-generation cephalosporin approved by the FDA in 1996 for treatment of complicated infections of urinary tract, skin and soft tissues, abdomen and lungs, as well as febrile neutropenia^1^. Among its side effects, the most concerning is an encephalopathy, initially presenting with impaired vigilance and evolving towards convulsions, coma, and status epilepticus^2^. Cefepime clearance is mainly renal and its plasma concentration can greatly differ between patients: upon intermittent IV administration, peak concentrations may vary four-fold whilst trough concentrations may vary up to forty-fold^3^.

Intensive care patients are especially prone to pharmacokinetic variability, due to distribution volume or clearance alterations elicited by acute kidney injury, sepsis, protein loss, or hyper-hydration^4, 5^. Consequently, they are at greater risk of experiencing drug toxicity or treatment failure^6^.

Previous studies have explored the pharmacokinetics/pharmacodynamics (PK/PD) targets for cefepime efficacy^3, 9, 10^. According to a simulation study, for patients with intermediate-severity illness (modified APACHE score between 9 and 22), 68% of free time above the minimum inhibitory concentration (*f* T>MIC) may be enough to improve the probability of survival^11^. Nonetheless, the European Society for Microbiology and Infectious Diseases recommend to achieve a 100% *f* T>MIC for all critically-ill adult patients treated with beta-lactams^12^. A recent meta-analysis showed that more “aggressive” beta-lactam therapy, with a target of 100% *f* T >4×MIC, yields higher clinical cure rates and lower risk of resistance development^13^.

From the safety point of view, a definite cut-off for cefepime through concentration has not been consensually established yet. Previous studies suggested cut-offs for neurotoxicity ranging between 16 and 49mg/L^14–18^.

Antimicrobial therapy in intensive care could benefit from model-informed precision dosing (MIPD): a personalized dosing advice based on therapeutic drug monitoring (TDM) and computation of individual PK parameters and dosage requirements, most often via a Bayesian PK approach^7, 8^. This approach showed significant clinical benefits for non-critically ill patients treated with vancomycin^19, 20^. In a randomized trial performed in critically ill patients, MIPD of beta-lactam and ciprofloxacin did not improve target attainment nor length of stay^21^, but further comments highlighted suboptimal performance of implemented models^22, 23^ and insufficient focus on high risk patients^23^. Of note, cefepime was not administered in this study.

Several cefepime population pharmacokinetics models have been developed and used to simulate the probability of target attainment^24–27^, but applications of such models for MIPD are still scarce. The objective of the present study was to assess the effect of MIPD on the cefepime concentration target attainment in patients admitted to the intensive care unit.

## Materials and methods

### Study design

We performed a monocentric, retrospective, before‒after study. The study protocol was approved by the Scientific and Ethical Committee of the *Hospices Civils de Lyon* and indexed with the study number 24-558. The statistical analysis plan was reviewed by internal peer reviewers but was not uploaded on an external repository. To ensure the completeness and the coherence of the present report, we followed the STROBE checklist for reporting of observational studies^28^ (see Supplementary Material).

All patients aged 18 years or more who were admitted to the intensive care unit of the *Centre Hospitalier Lyon Sud* and underwent at least one cefepime plasma concentration measurement between 01/01/2021 and 31/12/2024 were screened for the inclusion. Only patients with at least two cefepime plasma trough concentrations measured on separate days, as retrieved from the electronic record, were retrospectively included.

We did not perform a preliminary assessment of the number of subjects needed for inclusion, since we cannot find suitable preliminary data to base our simulations on. Instead, we adopted a pragmatical approach to balance the distribution of the main binary predictor (the MIPD) across our sample. Since the quasi-experimental intervention of interest was deployed in our hospital on 15/06/2023, the inclusion timespan was based on the hypothesis that realization of plasma cefepime dosages was constant over time.

### Quasi-experimental intervention

Prior to June 2023, cefepime initial dosage, TDM request and subsequent dosage adjustment were managed at the full discretion of ICU physicians. Of note, dosage adjustment was empirical, rather than being guided by pharmacokinetic modeling or other quantitative approaches.

Cefepime MIPD was routinely introduced in all the *Centre Hospitalier Lyon Sud’s* ICU units on 15/06/2023. At that time, a default prescription bundle of cefepime administration and blood draw for peak and through concentration measurements was implemented in the physician’s user interface of IntelliSpace Critical Care and Anesthesia (ICCA, *Koninklijke Philips N.V., Amsterdam, The Netherlands*). On-call physician was free to choose the initial cefepime dosing regimen and to modify the blood draw schedule. Pharmacy department was notified of all cefepime bundle prescription in ICU throughout a daily automated query on the electronic patient records run by ICCA. Upon validation of the TDM result by the pharmacotoxicology department, the BestDose^TM^ software (*LAPBK, University of Southern Californa, Los Angeles, CA, USA*) was run to estimate individual pharmacokinetic parameters and perform dosing simulations, based on one or two measured concentrations (a through alone or a couple of peak and trough). Two nonparametric population pharmacokinetic models were used^27, 29^. Both of them were made by two compartments and included an effect of renal function on cefepime elimination. For each patient and each measurement occasion, the choice between the two models was guided by the goodness-of-fit of the prediction. Then, a dosage adaptation was suggested to the ICU on-call physician, who kept the final decision on dose adjustment nonetheless. Dosing suggestions were provided within 12 hours since the validation of TDM result. As for the TDM service, the MIPD service was available from Monday to Friday, from 8:30 am to 6:00 pm.

Before and after the routine MIPD protocol introduction, in the present study cefepime was administered as a prolonged infusion of 3h, whatever the dose amount and dosing interval.

### Drug concentration measurement

The total plasma cefepime concentration was determined using high performance liquid chromatography (LC) with high-resolution mass spectrometer (HRMS) detection using a Q-Exactive Focus Orbitrap (*ThermoFisher Scientific, Sunnyvale, CA, USA)*. Sample preparation was based on a simple protein precipitation with methanol-HCOOH 0,1%. Chromatographic separation was performed on an Accucore C18 column 2.1 × 100 mm, 2.6 µm (*ThermoFisher Scientific, Sunnyvale, CA, USA*) using a binary elution gradient program with NH4HCO2 2mM buffer-HCOOH 0,1% and acetonitrile-HCOOH 0,1% as eluents. A volume of 10 µL was injected. A mobile phase flow rate of 0,5 mL/min was employed. Column oven and autosampler tray temperatures were 40°C and 10°C, respectively. The HRMS operated in full scan mode (from m/z 120 to 650, at a resolution of 70,000) after negative electrospray ionisation. Parameters for ionization were spray voltage 3.0 kV, nitrogen sheath gas pressure 60 arbitrary units, and auxiliary gas pressure 20 arbitrary units. The capillary temperature was 380 °C. Hardware control, data processing and treatment were carried out by Trace Finder v4.1 (*ThermoFisher Scientific, Sunnyvale, CA, USA*). The presence of the molecule was based on the mass of its parent ion with an accuracy of 5 ppm and on its retention time. The quantification was based on the principle of internal calibration using cefepim-D3 as internal standard with seven calibration points from 0,5 to 100 mg/L. Quadratic regression was selected using 1/x weighting factor. This method was validated over a range of 0,5 to 200 mg/L. Each batch of patient analysis included low- and high-quality controls respectively at 10 mg/L and 50 mg/L. The intra- and inter-assay variability of the quality controls was lower than 10% at both levels. This analytical method is routinely available at *Hospices Civils de Lyon* and is validated according to national French accreditation recommendations in medical biology (*Comité Français d’Accréditation*, *COFRAC*).

### Data collection and management

The database of the pharmacotoxicology department was first linked to the electronic patient record throughout a unique patient identifier. The following information was retrospectively collected from patients’ files:

- Age, sex, weight, plasma creatinine, and performance of renal replacement therapy;
- Site of infection, bacterial species detected by microbiological culture or multiplex PCR, and antimicrobial susceptibility testing (AST) results;
- Cefepime dosing history, TDM sampling times and measured concentrations.

Data were stored on a secured Access database (*Microsoft, Redmond, WA, USA*) during the collection phase. Except for the AST results, for which the imputation rules are described below, there were no other missing data and no additional imputation procedures were applied.

### Study endpoint

The main endpoint of our analysis was the achievement of target concentration range for cefepime through plasma concentration. The upper limit of the therapeutic range was 20mg/L, conservatively defined accordingly to the previous safety studies. The lower limit was defined as follows:

- If a MIC was known at the time of concentration measurement, then the limit was set to four times the MIC, according to previous efficacy studies. In case of polymicrobial infections, the highest MIC was selected;
- Elsewhere, the lower limit was set to a conservative « default » value of 10mg/L. Such cut-off was defined to achieve at least a target of 100% *f* T >1×MIC, considering that cefepime free fraction (*f*) is 80% and the highest epidemiological cut-off value reported by EUCAST is 8mg/L (for *P. aeruginosa*)^31^.

According to this ruleset, cases of isolated bacteria displaying intermediate resistance to cefepime (CMI ≥ 6mg/L) would generate a conflict, since 4×MIC ≥ 20mg/L. Nonetheless, such cases have been never encountered in the preliminary data review process, and no exceptions have been included in the analysis code.

If a patient had several concentration measurements, some could be interpreted with the « default » therapeutic range if results of AST were not available yet. Once the MIC was available, further concentration measurements were interpreted with the « personalized » therapeutic range. This approach aimed to reflect the clinical reasoning of practitioners who manually interpret both the AST and concentration measurements for their patients. For patients with multiple infectious episodes treated with cefepime, different targets may have been considered based on the pathogen(s) and their MIC, if these episodes were separated by at least one week without any antibiotic treatment.

### Statistical analysis

Data was described with absolute counts (and proportions), or with median (and quartiles 1 – 3) when appropriate. The first-type error rate tolerated for statistical analyses was 5%. Odds ratios and hazard ratios are reported with their 95% confidence interval.

We described the crude proportion of target attainment, as well as the proportion of under- and over-exposed patients, in the two groups. In addition, we described the probability distribution of observed concentrations.

The first through concentration was used only for descriptive purposes, but was censored in the further modelling phase, since it did not depend on the dosing optimization strategy.

We first assessed the proportional odds assumption across response categories to determine the suitability of an ordinal logit model. As this assumption was not met, we developed instead three separate binary logistic models: one for the odds of being under-exposed, one for the odds of being over-exposed, and one for the odds of being in the therapeutic range (versus being under- or over-exposed). Because of the design with repeated measures for each patient, we included a patient-centred random effect in the model formula. Models’ validity was assessed by comparison of observed and simulated throughs.

To answer whether MIPD helps attaining faster the therapeutic range, we modelled the rate of presenting the main endpoint. We started by censoring all the observations following the first therapeutic window attainment for each patient. Then, we computed the Kaplan-Meier survival estimator. We initially modelled the endpoint using a Cox proportional hazards model, which assumes proportionality of hazards over time. Subsequently, we developed a more flexible model for the event rate by modelling it as a cubic natural spline function of time with three knots, stratified by MIPD group.

The main predictor of our analysis was the quasi-experimental group (that is, the indicator of the MIPD group versus the control group). Other clinical variables were tested during the modelling phase, and final multivariate model selection was based on the deviance as well as on clinical significance. In particular, we expected a possible confounding effect of the dosing strategy and of creatinine clearance.

Logit and hazard models’ equations are provided in the Supplementary Material. Statistical analyses were implemented with R 4.4.3 (*GNU license*). Logistic models’ likelihood was maximized using *lme4*^32^, and assumptions were tested by simulation with *DHARMa*^33^. Rate models’ likelihood was maximized using *survPen*^34^.

### Sensitivity analyses

We performed several sensitivity analyses to assess the robustness of the results and to account for uncertainty in PK measurements and variability in PK/PD targets.

In sensitivity analysis 1, which we applied to both the odds and the rate modelling, we censored trough concentrations with sampling time more than 2 hours prior to the next administration, as such values are overestimates of the real trough. Of note, such information was not considered by clinicians for dose adjustment prior to routine MIPD implementation, while it was correctly considered in the MIPD period.

Additional sensitivity analyses were conducted for the odds modelling. In sensitivity analysis 2, follow-up was censored at one week for all patients to evaluate the potential bias introduced by longer follow-up, which may be associated with greater severity or inter-observation variability. In sensitivity analysis 3, we censored the patients for which MIPD was performed before the 15/06/2023, as well as those included afterwards and for which MIPD was not performed. By doing so, we wanted to assess the impact of the secular trend on our series. In sensitivity analysis 4, we fixed the lower margin of the therapeutic range at 10mg/L for all patients, regardless of MIC availability, to assess the impact of a dynamic endpoint definition. In sensitivity analysis 5, we fixed the lower therapeutic margin at 1×MIC for all patients, to evaluate the impact of using an alternative PK/PD target.

## Data and code availability

Anonymous data is available upon request to the corresponding author. R code used for data analysis is publicly available on https://osf.io/gtcdm/ under a CC-By Attribution 4.0 International license.

## Results

### Patients’ characteristics

Among the 512 patients screened, 281 were included in the study. The main reason for exclusion was that TDM was only performed once, and so we could not evaluate the effect of dose adjustment on cefepime exposure. 130 patients were included before the date of MIPD implementation in routine care, and 151 after. For 4 patients included in the «before» period, cefepime was dosed using MIPD, and these patients were analysed in the MIPD group. For 34 patients included in the «after» period, MIPD was not performed, and these patients were analysed in the control group. A total of 121 (43%) and 160 (57%) patients were analysed in the MIPD group and control group respectively. The study flow chart is shown in Figure 1.

**Figure 1:**
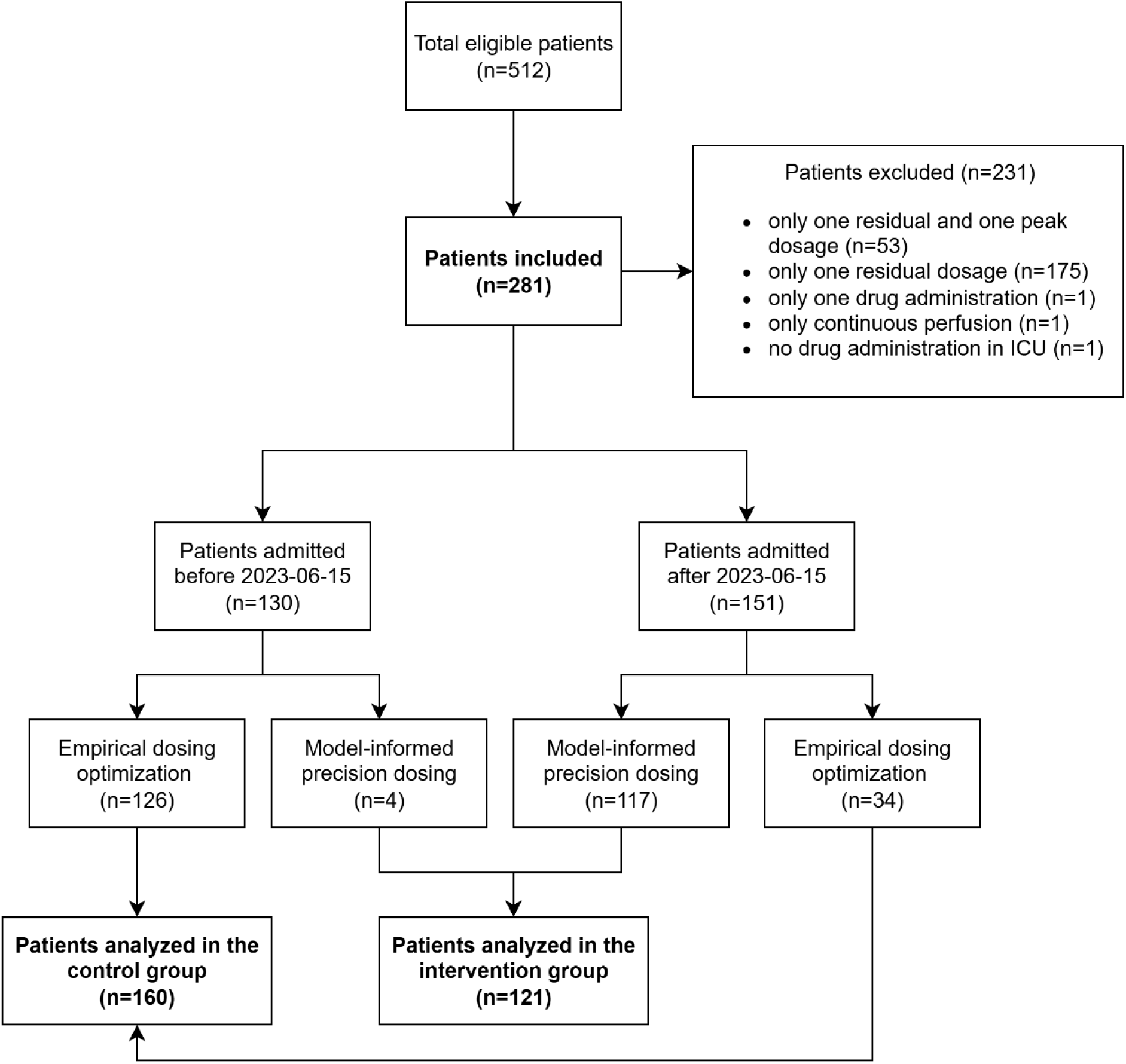
flowchart of patients inclusion and analysis group allocation in the present study.

Patients’ characteristics are shown in Table 1. In the full sample, male/female sex ratio was 2.09, age was 65 (Q1 – Q3: 56 – 74) years, weight was 76 (65 – 90) kg, and follow-up duration was 3 (1.9 – 4.9) days. These characteristics were consistent between groups. Creatinine clearance was slightly imbalanced between groups, being 100 (95 – 104) mL/min in the MIPD group and 86 (81 – 91) mL/min in the control group. Accordingly, renal replacement therapy was used more frequently in the control group (Table 1).

**Table 1:** characteristics of patients included in the present study.

|  | **Overall**<br>N = 281<sup>1</sup> | **MIPD**<br>N = 121<sup>1</sup> | **Control**<br>N = 160<sup>1</sup> |
| --- | --- | --- | --- |
| Male sex | 190 (68%) | 85 (70%) | 105 (66%) |
| Age (years) | 65 (56 – 74) | 64 (53 – 74) | 66 (58 – 74) |
| Weight (kg) | 76 (65 – 90) | 74 (64 – 87) | 77 (66 – 91) |
| Plasma creatinine (µmol/L) | 67 (65 – 71) | 63 (60 – 66) | 71 (67 – 76) |
| CrCl (mL/min) | 95 (88 – 100) | 100 (95 – 104) | 86 (81 – 91) |
| RRT – of which: | 30 (10%) | 6 (5%) | 24 (15%) |
| IHD | 10 (3%) | 0 | 10 (6%) |
| CVVHD | 20 (7%) | 6 (5%) | 14 (9%) |
| Follow-up duration (days) | 3 (1.9 – 4.9) | 3 (2 – 4.5) | 3 (1.5 – 5) |
| Through concentration<br>(all samples, n) | 728 | 338 | 390 |
| Through concentration<br>(all samples, mg/L) | 23 (13 – 42) | 21 (12 – 37) | 25 (13 – 44) |
| Through concentration<br>(first samples, n) | 267 | 114 | 153 |
| Through concentration<br>(first samples, mg/L) | 27 (15 – 50) | 27 (13 – 47) | 29 (16 – 52) |
| Through concentration<br>(further samples, n) | 461 | 224 | 237 |
| Through concentration<br>(further samples, mg/L) | 21 (12 – 36) | 19 (11 – 32) | 22 (13 – 42) |
| Peak concentration (n) | 264 | 170 | 94 |
| Peak concentration (mg/L) | 55 (41 – 77) | 54 (42 – 73) | 56 (38 – 82) |

<sup>1</sup>n (%); Median (Q1 – Q3).
Abbreviations. CrCl: creatinine clearance estimated via Cockcroft-Gault formula. CVVHD: continuous veno-venous haemodialysis. MIPD: model-informed precision dosing. IHD: intermittent haemodialysis. RRT: renal replacement therapy.

A total of 301 infectious episodes were recorded in the full sample. The most common infection type was pneumonia (n=239, 79%). Among abdominal infections (n=29, 9.6%), the most common infection was peritonitis (n=16, 5%). These proportions were consistent between groups (Table S1). 418 bacteria were isolated, of which 409 (97.8%) were Gram-negative bacteria. The most common pathogen was *P. aeruginosa* (n=191, 45.7%), followed by *E. coli* (n=46, 11%) (Table S2).

75 AST were performed in the full sample (17.9% of isolated bacteria), and the median MIC was 1 (0.12 – 2) mg/L. In the MIPD group, 37 AST were available (20.7% of isolated bacteria in that group) and MIC was 0.25 (0.12 – 2) mg/L. In the control group, 38 AST (15.8%) were available and MIC was 2 (0.75 – 2) mg/L.

After adjustment for infectious episode and timing of MIC measurement, the lower limit of the « personalized » therapeutic range was 10 mg/L in 85% of cases in both groups. The second most common lower limit was 8mg/L, occurring in 4.7% of cases in the MIPD group and 5.9% in the control group (this resulted from the multiplication 4 × a known MIC of 2mg/L).

### Concentration measurements

A total of 728 cefepime trough concentrations and 264 peak concentrations were measured in the full sample. Peak concentrations were 54 (42 – 73) mg/L in the MIPD group and 56 (38 – 82) mg/L in the control group. Through concentrations in the second and further measurements were 19 (11 – 32) mg/L in the MIPD group and 22 (13 – 42) mg/L in the control group (Table 1). Those measured in the MIPD group showed a similar median but a slightly lower variability than those in the control group (Figure S1).

When considering only the through concentrations in the second and further measurements, the proportion of patients in the therapeutic range was 35% and 28% in the MIPD and control group, respectively (Table 2).

**Table 2:** proportions and odds ratios of target attainment of cefepime through concentrations (first samples for each cefepime course were not included).

|  | **Overall**<br>N = 461<sup>1</sup> | **MIPD**<br>N = 224<sup>1</sup> | **Control**<br>N = 237<sup>1</sup> | **Crude OR<sup>2</sup>** | **Adjusted OR<sup>2,3</sup>** |
| --- | --- | --- | --- | --- | --- |
| Under-exposure | 81 (18%) | 45 (20%) | 36 (15%) | 1.71 [0.76 – 3.86] |  |
| Target attainment | 146 (32%) | 79 (35%) | 67 (28%) | 1.44 [0.9 – 2.31] | 1.4 [0.88 – 2.22] |
| Over-exposure | 234 (51%) | 100 (45%) | 134 (57%) | 0.52 [0.3 – 0.88] | 0.58 [0.35 – 0.96] |

<sup>1</sup>n (%). <sup>2</sup> odds ratio with 95% confidence interval within square brackets. <sup>3</sup> Estimated with a multivariate logistic mixed-effect model adjusted on MIPD and creatinine clearance. Model for under-exposure did not converge.
Abbreviations. MIPD: model-informed precision dosing.

Starting from the third concentration measurement, patients in the MIPD group had a lower variability of trough concentrations than control patients (Figure 2), as well as a higher probability of target attainment (Figure 3).

**Figure 2:**
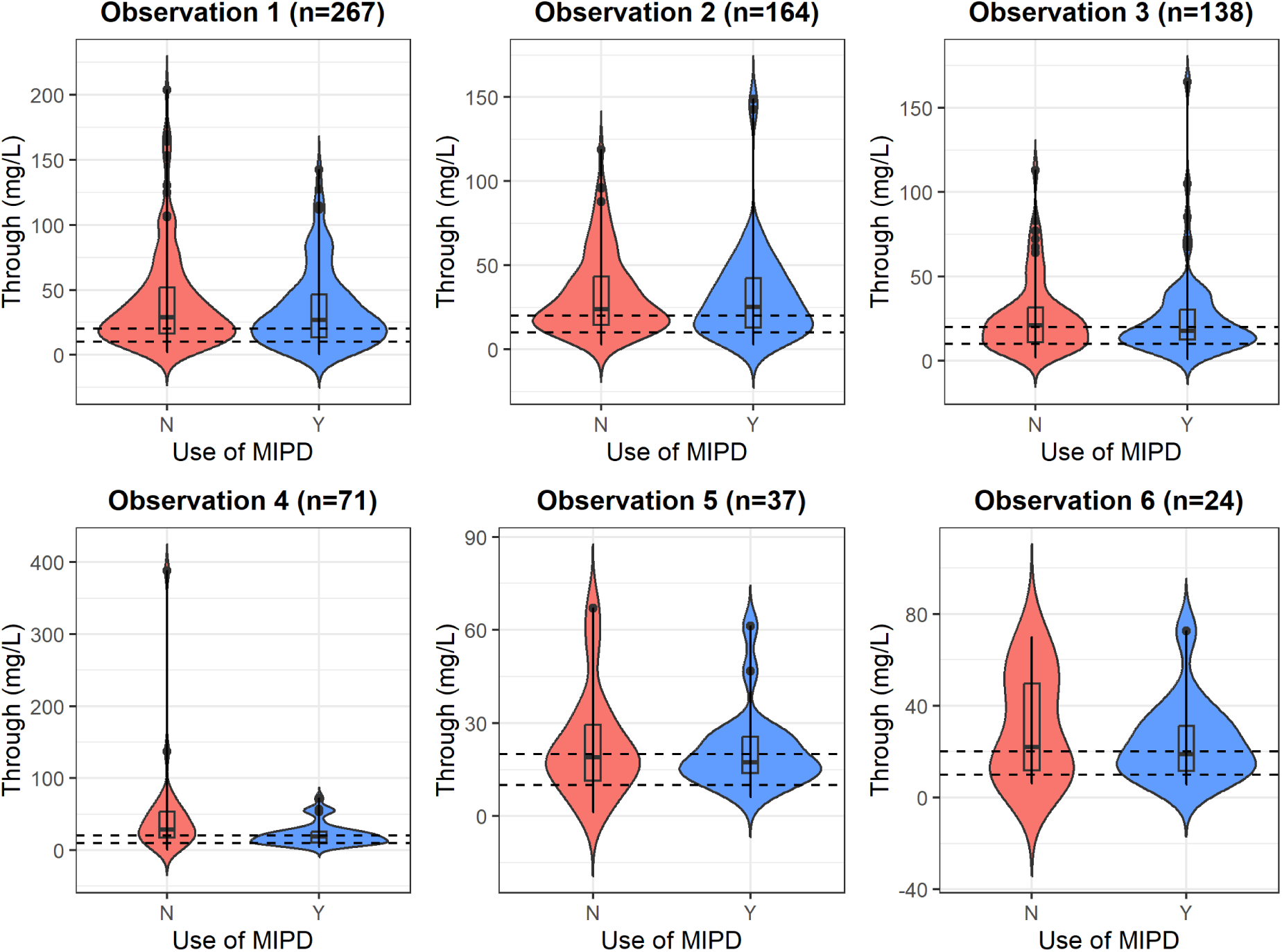
distribution of trough concentrations, stratified by group and observation. Blue: MIPD group. Red: control group. Dashed lines: default therapeutic window comprised between 10mg/L and 20mg/L. Observations later than 6 have been censored due to sparsity of data.

**Figure 3:**
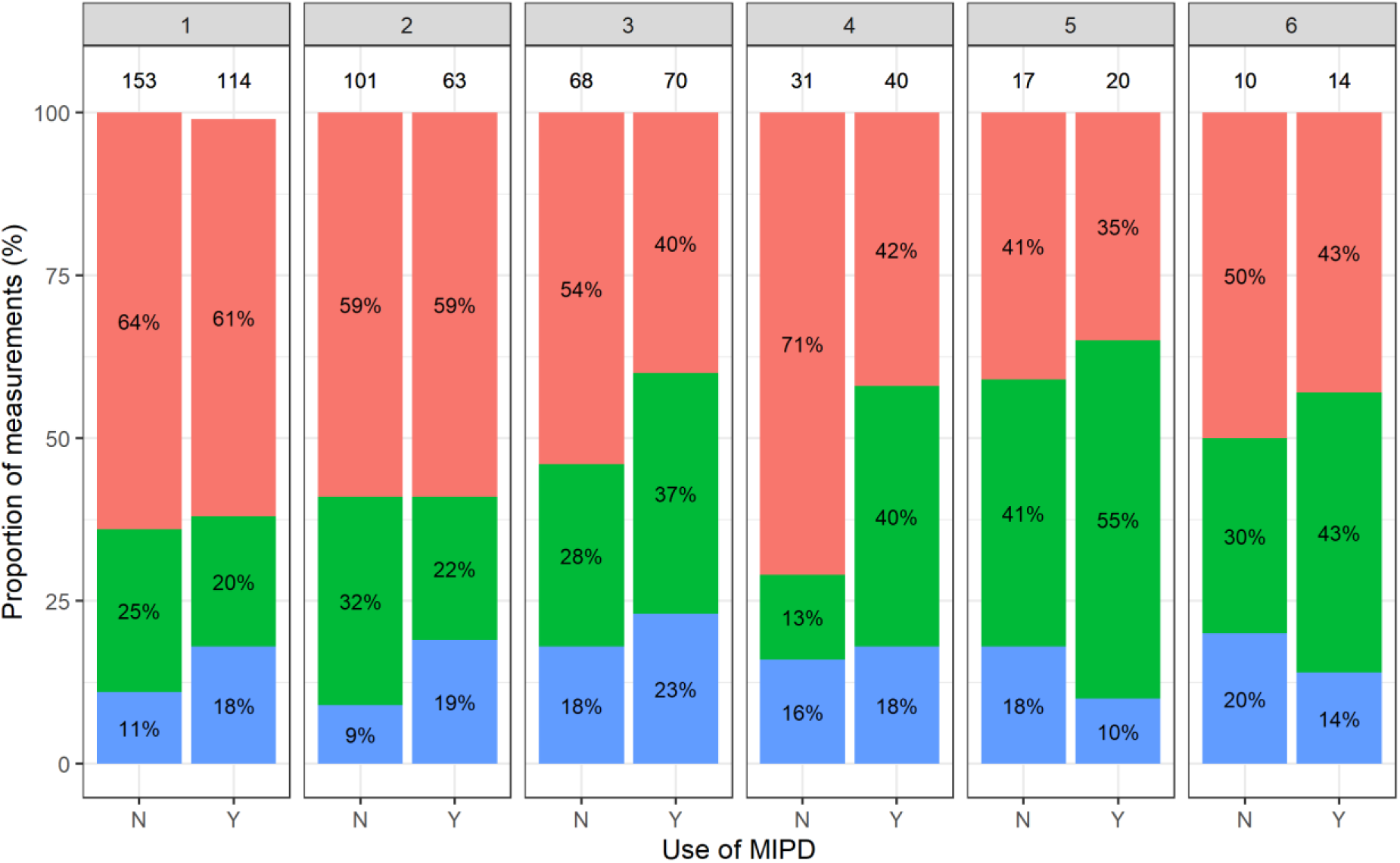
proportion of measured through concentration in the therapeutic window, stratified by group and occasion. Blue: through concentration under 10 mg/L. Green: through concentration within the therapeutic target range. Red: through concentration above 20mg/L. Absolute patient counts for each group and observation are displayed above the stacked bars. Percentages of target attainment are showed inside the bars. Observations later than 6 have been censored due to sparsity of data.

Dosing schemes changes in the MIPD group were more skewed toward a dosing reduction, whereas physicians of the control group increased dosing more often. Regarding the dose adjustment in the MIPD group, cefepime dose was left unchanged in 65% of cases, reduced in 27%, augmented in 8% of cases, whereas in the control group, it was left unchanged in 61%, reduced in 24%, and augmented in 14% (Figure S3).

### Modelling of odds

In univariate analysis, MIPD was associated with higher odds of attaining the therapeutic range as well as higher odds of being under-exposed, but these associations did not reach statistical significance. On the other hand, the association between MIPD and lower odds of over-exposure was statistically significant (OR 0.52, IC 95% 0.30 – 0.88).

These results were confirmed by multivariate analysis (adjusted on the effect of creatinine clearance). MIPD was still positively associated with higher odds of attaining the therapeutic range (aOR 1.40, IC 95% 0.88 – 2.22) and lower odds of being over-exposed (aOR 0.58, IC 95% 0.35 – 0.96, Table 2).

An increase of creatinine clearance (doubling from the reference value of 122 mL/min) was associated with higher odds of being under-exposed (aOR 1.94, 95% CI 1.27 – 2.97) or correctly exposed (aOR 1.41, 95% CI 1.14 – 1.74), and lower odds of being over-exposed (aOR 0.41, 95% CI 0.30 – 0.56). For instance, a patient with a creatinine clearance of 60 mL/min (that is, a 49% reduction from the reference value) had (–49%) × (–59%) = 29% higher odds of being over-exposed, when compared with a patient with a creatinine clearance of 122 mL/min.

### Modelling of rate

A total of 112 patients (92%) in the MIPD group and 145 patients (90%) in the control group did not achieve a satisfactory through concentration at their first control and were considered « at risk » (Table 3). The log-rank test showed no statistically significant difference between groups (Kaplan-Meier plot is showed in Figure 4, top panel). In the MIPD group, the peak rate of target attainment (0.21 patients/day) occurred twelve hours after the peak rate in the control group (Figure 4, mid panel). The predicted hazard ratio was not constant over time, being 1.10 (IC 95% 0.7 – 1.9) at day 1, 1.30 (IC 95% 0.9 – 1.9) at day 3 and 1.60 (IC 95% 0.9 – 2.9) at day 7 (Figure 4, bottom panel; Table 3).

**Figure 4:**
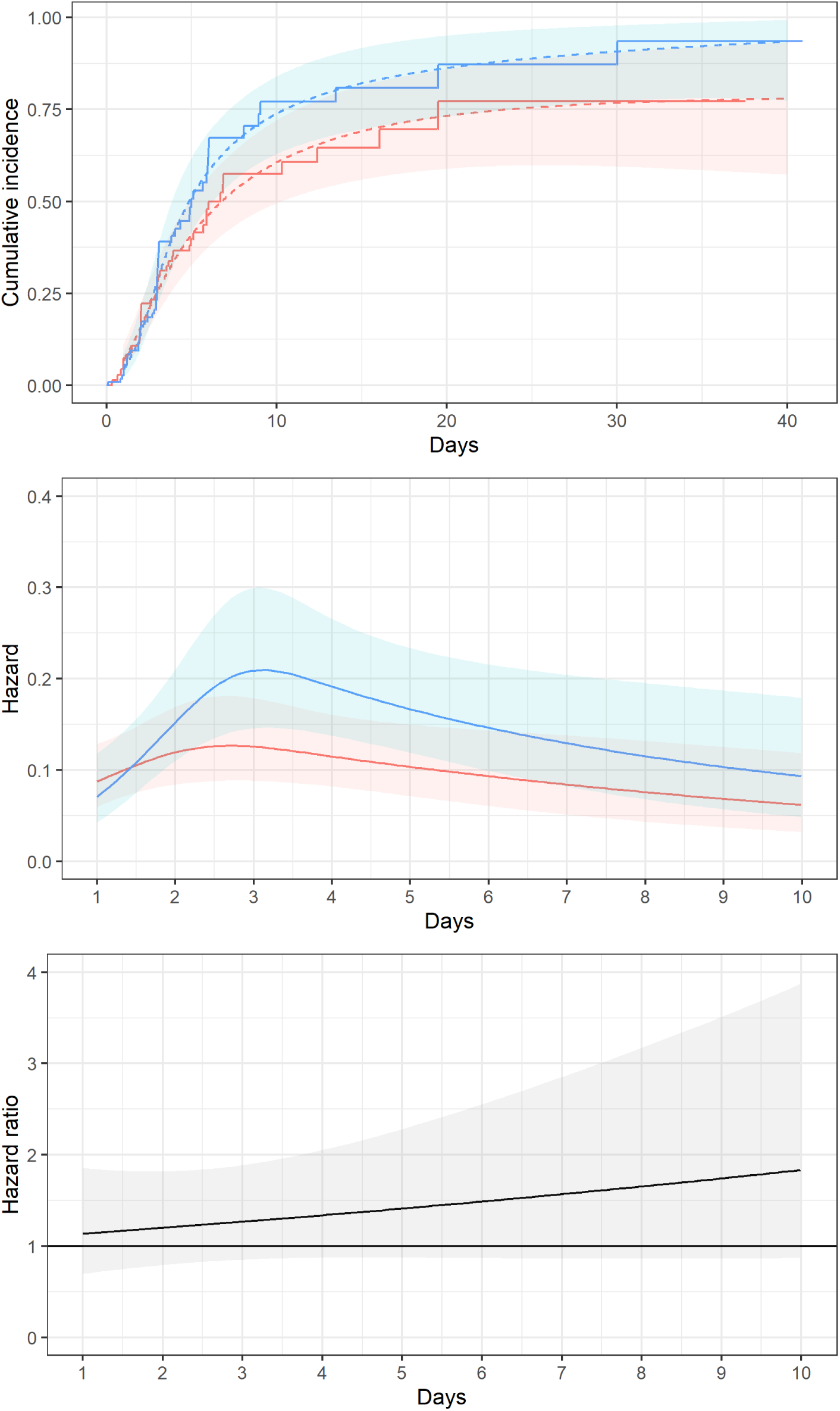
results of modelling of time to target attainment. Blue: MIPD group. Red: control group. **Top panel:** cumulative incidence of target attainment. Solid line: Kaplan-Meyer estimator. Dashed line: cumulative incidence predicted under the rate flexible model. Coloured area: 95% confidence interval of prediction. **Middle panel:** target attainment rate predicted by the flexible model. Coloured area: 95% confidence interval of prediction. **Bottom panel:** hazard ratio predicted under the rate flexible model. Grey area: 95% confidence interval of prediction. Prediction of hazard and hazard ratio later than day 10 are not shown, due to sparsity of input data.

**Table 3:** risk table for target attainment in the present study.

| Day |  | 0 | 1 | 2 | 3 | 4 | 5 | 6 | 7 | 10 | 20 | 30 |
| --- | --- | --- | --- | --- | --- | --- | --- | --- | --- | --- | --- | --- |
| MIPD | at risk<sup>1</sup> | 112 | 103 | 84 | 54 | 32 | 23 | 14 | 13 | 7 | 2 | 2 |
|  | events<sup>2</sup> | 0 | 3 | 16 | 31 | 37 | 42 | 48 | 49 | 52 | 54 | 54 |
| Control | at risk<sup>1</sup> | 145 | 122 | 97 | 67 | 44 | 36 | 23 | 17 | 15 | 3 | 2 |
|  | events<sup>2</sup> | 0 | 10 | 19 | 32 | 39 | 41 | 46 | 49 | 49 | 53 | 53 |
| HR<sup>3</sup> |  |  | 1.1<br>[0.7 - 1.9] | 1.2<br>[0.8 - 1.8] | 1.3<br>[0.9 - 1.9] | 1.4<br>[0.9 - 2.1] | 1.5<br>[0.9 - 2.3] | 1.5<br>[0.9 - 2.6] | 1.6<br>[0.9 - 2.9] | 1.8<br>[0.9 - 3.9] | 3.0<br>[0.5 – 17.4] | 4.8<br>[0.1 – 343] |

<sup>1</sup> Were considered «at risk» all the patients who had never had a cefepime through concentration within the target range yet.
<sup>2</sup> The considered «event» was the attainment of the target range of cefepime through concentration.
<sup>3</sup> Hazard ratio for target attainment predicted under the rate flexible model with 95% confidence interval within square brackets.
Abbreviations. MIPD: model-informed precision dosing.

### Sensitivity analyses

In sensitivity analysis 1, we censored 109 through concentrations sampled more than 2 hours before the following cefepime administration (58 in the control group and 51 in the MIPD group). As for the principal analysis, through concentrations from the second measurement onwards showed a similar median but lower variability in the MIPD group than the control. Such trend was greater from the third measurement onwards (data not shown). Proportion of patients attaining the target range increased from the third concentration measurement onwards (Figure S2). The effect size of MIPD on the adjusted odds of attaining the therapeutic range was 23% lower than in the primary analysis. On the other hand, MIPD effect on the adjusted odds of being over-exposed was only 8% lower (Table S3). These results are consistent with the primary odds analysis. The application of sensitivity analysis 1 to the hazard modelling yielded results consistent with the primary analysis as well, but with a smaller effect size on the hazard ratio (Table S4, Figure S4).

In sensitivity analysis 2, we censored 82 through concentrations (36 in the MIPD group and 46 in the control group) sampled after a follow-up time greater than 7 days. The impact of such analysis on the adjusted odds estimation was limited, and consistent with the primary odds analysis (Table S3).

In sensitivity analysis 3, we censored 4 patients for which MIPD was performed before the 15/06/2023, as well as 34 patients included afterwards and for which MIPD was not performed. The effect on the odds of being in the therapeutic range were increased by only 3%, and the odds of being over-exposed were unchanged (Table S4).

Sensitivity analysis 4 and 5 showed little to no impact on the estimations of the odds of target attainment (Table S4).

## Discussion

There is a growing interest about MIPD in antimicrobial therapy. Previous studies showed mixed results^20, 21^, raising the need to better identify which drugs are the best candidates and which populations of patients should benefit the most from this practice. Cefepime is a good candidate for TDM and MIPD, considering the established exposure-dependant neurotoxicity, and critically ill patients are obviously a population of interest. To our knowledge, this is the first evaluation of the impact of MIPD for cefepime in critically ill patients.

Our real-world data analysis demonstrates that model-informed precision dosing of cefepime in the intensive care unit is associated with lower odds of being over-exposed compared to empirical therapeutic drug monitoring-based dose adjustments. Since the through concentration over the 20mg/L threshold is a validated proxy for neurological toxicity^15^, we can infer that MIPD may help to prevent cefepime neurological toxicity in the intensive care unit.

When provided with model-informed advice by pharmacists, our ICU physicians reduced cefepime doses more often than they did in the empirical dose-adjustment period. Even though we did not test the statistical significance of this observation, this fact is directly associated with the effect of MIPD on preventing over-exposure.

In our study, MIPD has a positive effect on the odds as well as on the rate of target attainment. In other words, when MIPD was deployed, target was attained by more patients, and it was attained faster, than with empirical dose adjustments. Nonetheless, such results did not reach statistical significance, and they should be interpreted cautiously. The proportion of target attainment in our MIPD group was low (37% on the third observation, and 55% on the fifth observation), despite repeated concentration measurements and subsequent dosing adaptations. Our result agrees with DOLPHIN trial’s results^21^, which reported a 55% target attainment at day one and a 59% target attainment at day 3. Taken together, these results should raise questions on the performance of predictive pharmacokinetics models currently implemented in MIPD, especially regarding their robustness to intra-individual pharmacokinetic variability.

The descriptive analysis of cefepime concentrations measured in our study suggests a slightly lower variability of concentrations in the MIPD group. This fact could result from the protective effect of MIPD against over-exposures to cefepime. Nonetheless, the size of this effect seems modest and its statistical significance has not been evaluated. Further research should assess more precisely the difference in error terms of non-linear mixed-effects models to answer to the question whether MIPD reduces the inter- and intra-individual variability.

The overall results of our sensitivity analyses are coherent with our primary analysis. The effect reduction observed in sensitivity analysis 1 (censoring of through concentration sampled more than 2 hours from next administration) offer some insight on the risk of residual measurement bias of the primary analysis. The effect reduction observed in sensitivity analysis 2 (censoring of follow-up longer than 7 days) suggests that MIPD could be more useful for patients who need multiple concentration measurements, because of a long cefepime course or because of a great intra-individual variability of concentration. Results of sensitivity analysis 3 (censoring of patients having the intervention of the other group) rule out the risk of secular trend bias in our primary analysis. Results of sensitivity analysis 4 and 5 (choice of a fixed lower margin of the therapeutic range) rule out the risk of misclassification bias due to our dynamic therapeutic range definition.

The added value of TDM compared to fixed dosing schemes has been already established. Some randomized controlled trials (RCTs) comparing beta-lactams TDM versus standard dosing strategies did find a beneficial effect of the intervention on the probability of target attainment^35, 36^, but no effect on clinical endpoints^36^. This result is confirmed by a recent meta-analysis which included both RCTs and observational studies^37^.

Taken together, our results suggest that MIPD provides additional safety in cefepime administration, compared to the TDM-based empirical dose optimization. Nonetheless, our work failed to demonstrate a significant association between MIPD and target attainment, as well as previous RCTs comparing MIPD versus TDM-based empirical dosing optimization strategies. The “Right dose, right now” trial, which was stopped early due to the COVID-19 pandemics, failed to demonstrate a benefit of beta-lactams MIPD on both PK/PD target attainments and clinical outcomes^38^. The DOLPHIN trial also failed to demonstrate a beneficial effect of MIPD on the intensive care unit length of stay^21^, even though its results could be biased toward a lesser intervention effect size estimation due to some flaws in the implemented models and in patients selection^22, 23^.

A possible explanation of the lack of efficacy of MIPD (and TDM) in studies on clinical endpoints may be the excessive delay to obtain TDM and MIPD results. Indeed, it is early PK/PD target attainment that is critical to optimize clinical efficacy. The BLAST 1 observational study showed that the target attainment within the first 48 hours of treatment is significantly associated with better clinical outcomes such as mortality and length of stay^39^. For instance, the quasi-experimental study by Hall et al. showed a significant clinical benefit of vancomycin MIPD compared to TDM-based optimization, and their model-based AUC monitoring was performed early in the workup, before the attainment of steady state concentration^20^. From this point of view, our estimation of MIPD effect on target attainment hazard ratio should be further developed. The effect size estimations provided by the present study (HR 1.2 at 48 hours) may be a valuable insight to be used in power simulations for future interventional studies.

We must point out several limitations of the present work. Our main endpoint (target concentration attainment) is a surrogate endpoint, and further prospective, controlled studies would be necessary to confirm the value of cefepime MIPD to optimize clinical efficacy and safety. In addition, we could assess renal function only through the Cockroft-Gault estimation of creatinine clearance, rather than the urine/plasma creatinine ratio, which is a less biased estimator of glomerular filtration rate in intensive care patients^40, 41^. These two elements may have introduced some measurement biases in our analysis.

Since the inclusion in our retrospective study was not stratified, we observed too few patients with RRT (and, moreover, they were not fairly distributed among groups), and we could not adjust for its potential confounding effect. Thus, our results may not be generalizable to this sub-population, and further targeted studies should be performed. In addition, the likelihood maximization algorithm of the multivariate logistic model for the odds of under-exposure did not converge, probably because of the scarcity of under-exposed patients. Therefore, we could not assess the confounding effect of creatinine clearance on such endpoint.

Some biases could have been introduced by the observational nature of our study. First, it should be noted that, in our hospital, TDM and MIPD are not available 24/7. Thus, physicians in charge of patients of the experimental group may have performed some empirical dosing optimizations. This fact could have introduced a treatment crossover effect in our data and ultimately lead to a lesser estimation of the MIPD effect. Furthermore, according to our hospital’s routine care protocols, physicians are always free to choose to not follow the pharmacy’s dosing advice. We did not measure the proportion of agreement between the dosing advice provided by pharmacists and the actual dosing adjustment made by physicians. Such methodological choice has the advantage to be comparable to the « intention to treat » principle of interventional research, but it may provide a lesser estimation of intervention effect.

Some measured through concentrations were surprisingly high, even in patients without renal impairment. This could be due to some pre-analytical errors, such as errors in the blood draw procedure or wrong labelling of sample tubes. Since it was impossible for us to retrospectively detect this type of errors, we could not censor these measurements, which introduced some noise in our data. Nonetheless, we expect such bias to be non-differential between groups, and thus not to impact the interpretation of our primary results.

Finally, the lack of preliminary power calculations may have limited our inference, as showed by the large confidence intervals of our estimations.

In conclusion, our real-world retrospective study suggests that MIPD in the intensive care unit reduces the risk of over-exposure to cefepime, and may be beneficial both on the odds and on the rate of PK/PD target attainment.

## Data Availability

Anonymous data is available upon request to the corresponding author. R code used for data analysis is publicly available on the Open Science Framework under a CC-By Attribution 4.0 International license.

https://osf.io/gtcdm/

## Contributions (CRediT taxonomy)

Conceptualization: A.F. and S.G.; Data curation: G.B. and S.C.; Formal analysis: G.B.; Investigation: G.B., F.W., S.P., R.G.; Methodology: G.B.; Project administration: A.F.; Resources: A.F.; Software: G.B.; Supervision: A.F. and S.G; Validation: S.G.; Visualization: G.B.; Writing – original draft: G.B.; Writing – review & editing: A.F., G.B., R.G., S.C., S.G., S.P.

