## Supplementary material for "Evaluation of model–informed precision dosing of cefepime in critically ill patients: a French before– after study"

### Table of contents

| | |
|---|---|
| Table S1 | page 2 |
| Table S2 | page 3 |
| Table S3 | page 4 |
| Table S4 | page 5 |
| Figure S1 | page 6 |
| Figure S2 | page 6 |
| Figure S3 | page 7 |
| Figure S4 | page 8 |
| STROBE checklist | page 9 |

**Table S1:** Types of infection.

| | **Overall**<br>N = 301[1] | **MIPD**<br>N = 172[1] | **Control**<br>N = 129[1] |
|---|---|---|---|
| Abdominal infection | 29 (9.6%) | 17 (9.9%) | 12 (9.3%) |
| Febrile neutropenia | 1 (0.3%) | 1 (0.6%) | 0 (0%) |
| Osteitis | 2 (0.7%) | 1 (0.6%) | 1 (0.8%) |
| Pericarditis | 1 (0.3%) | 1 (0.6%) | 0 (0%) |
| Pneumonia | 239 (79%) | 142 (83%) | 97 (75%) |
| Septic shock | 4 (1.3%) | 1 (0.6%) | 3 (2.3%) |
| SSTI | 13 (4.3%) | 3 (1.7%) | 10 (7.8%) |
| Urinary infection | 12 (4.0%) | 6 (3.5%) | 6 (4.7%) |

[1]n (%).

Abbreviations. SSTI: skin and soft tissues infection

**Table S2:** bacterial isolates.

| | **Overall** N = 418[1] | **MIPD** N = 178[1] | **Control** N = 240[1] |
|---|---|---|---|
| *Achromobacter xylosoxidans* | 2 (0.5%) | 1 (0.6%) | 1 (0.4%) |
| *Acinetobacter baumannii* | 2 (0.5%) | 0 (0%) | 2 (0.8%) |
| *Buttiauxella agrestis* | 1 (0.2%) | 1 (0.6%) | 0 (0%) |
| *Citrobacter braakii* | 6 (1.4%) | 3 (1.7%) | 3 (1.3%) |
| *Citrobacter freundii* | 6 (1.4%) | 4 (2.2%) | 2 (0.8%) |
| *Citrobacter koseri* | 3 (0.7%) | 1 (0.6%) | 2 (0.8%) |
| *Corynebacterium striatium* | 1 (0.2%) | 0 (0%) | 1 (0.4%) |
| *Enterobacter cloacae* | 26 (6.2%) | 16 (9.0%) | 10 (4.2%) |
| *Enterobacter ludwigii* | 1 (0.2%) | 1 (0.6%) | 0 (0%) |
| *Escherichia coli* | 46 (11%) | 16 (9.0%) | 30 (13%) |
| *Haemophilus influentiae* | 17 (4.1%) | 5 (2.8%) | 12 (5.0%) |
| *Hafnia alvei* | 1 (0.2%) | 1 (0.6%) | 0 (0%) |
| *Klebsiella aerogenes* | 24 (5.7%) | 12 (6.7%) | 12 (5.0%) |
| *Klebsiella oxytoca* | 12 (2.9%) | 7 (3.9%) | 5 (2.1%) |
| *Klebsiella pneumoniae* | 34 (8.1%) | 16 (9.0%) | 18 (7.5%) |
| *Moraxella catarrhalis* | 6 (1.4%) | 1 (0.6%) | 5 (2.1%) |
| *Morganella morganii* | 5 (1.2%) | 2 (1.1%) | 3 (1.3%) |
| Other Gram negative | 1 (0.2%) | 0 (0%) | 1 (0.4%) |
| *Proteus mirabilis* | 7 (1.7%) | 3 (1.7%) | 4 (1.7%) |
| *Proteus vulgaris* | 3 (0.7%) | 2 (1.1%) | 1 (0.4%) |
| *Pseudomonas aeruginosa* | 189 (45%) | 75 (42%) | 114 (48%) |
| *Pseudomonas aeruginosa* (mucoid) | 2 (0.5%) | 2 (1.1%) | 0 (0%) |
| *Serratia liquefaciens* | 1 (0.2%) | 0 (0%) | 1 (0.4%) |
| *Serratia marcescens* | 12 (2.9%) | 7 (3.9%) | 5 (2.1%) |
| *Serratia odorifera* | 2 (0.5%) | 1 (0.6%) | 1 (0.4%) |
| *Staphylococcus aureus* | 3 (0.7%) | 1 (0.6%) | 2 (0.8%) |
| *Streptococcus agalactiae* | 1 (0.2%) | 0 (0%) | 1 (0.4%) |
| *Streptococcus anginosus* | 1 (0.2%) | 0 (0%) | 1 (0.4%) |
| *Streptococcus constellatus* | 1 (0.2%) | 0 (0%) | 1 (0.4%) |
| *Streptococcus mitis* | 1 (0.2%) | 0 (0%) | 1 (0.4%) |
| *Streptococcus mutans* | 1 (0.2%) | 0 (0%) | 1 (0.4%) |

[1]n (%); Median (Q1, Q3)

**Table S3:** sensitivity analyses of the odds modelling. Odds ratios (OR) are reported with their 95% confidence interval within square brackets.

| | Target attainment | Under-exposure | Over-exposure |
|---|---|---|---|
| **Primary analysis** (crude OR) | 1.44<br>[0.9 - 2.31] | 1.71<br>[0.76 - 3.86] | 0.52<br>[0.3 - 0.88] |
| **Primary analysis** (adjusted OR) | 1.4<br>[0.88 - 2.22] | | 0.58<br>[0.35 - 0.96] |
| **Sensitivity analysis 1**<br>(only throughs < 2 hours) | 1.17<br>[0.72 - 1.91] | 1.57<br>[0.27 - 9.12] | 0.66<br>[0.37 - 1.15] |
| **Sensitivity analysis 2**<br>(only follow-up < 7 days) | 1.12<br>[0.67 - 1.87] | 1.53<br>[0.21 - 10.85] | 0.68<br>[0.39 - 1.18] |
| **Sensitivity analysis 3**<br>(only patients respecting intervention group) | 1.47<br>[0.89 - 2.43] | 1.56<br>[0.26 - 9.22] | 0.52<br>[0.29 - 0.92] |
| **Sensitivity analysis 4**<br>(lower therapeutic margin 10mg/L) | 1.44<br>[0.93 - 2.22] | 1.46<br>[0.69 - 3.12] | |
| **Sensitivity analysis 5**<br>(lower therapeutic margin 1×MIC) | 1.38<br>[0.9 - 2.14] | 1.35<br>[0.23 - 7.99] | |

Abbreviations. MIC: minimum inhibitory concentration.

**Table S4:** sensitivity analysis 1 of target attainment rate modelling. Through concentrations measured on blood samples drew at least 2 hours before the real pharmacokinetic through point have been censored. Hazard ratio (HR) and its 95% confidence interval were predicted under the flexible model of rate.

| Days | HR | 95% CI |
|---|---|---|
| 1 | 1.0 | 0.6 - 1.7 |
| 2 | 1.0 | 0.7 - 1.6 |
| 3 | 1.0 | 0.7 - 1.6 |
| 4 | 1.0 | 0.7 - 1.6 |
| 5 | 1.1 | 0.7 - 1.7 |
| 6 | 1.1 | 0.7 - 1.7 |
| 7 | 1.1 | 0.6 - 1.9 |
| 10 | 1.2 | 0.6 - 2.3 |
| 20 | 2.0 | 0.5 - 7.9 |
| 30 | 4.4 | 0.3 - 66.4 |

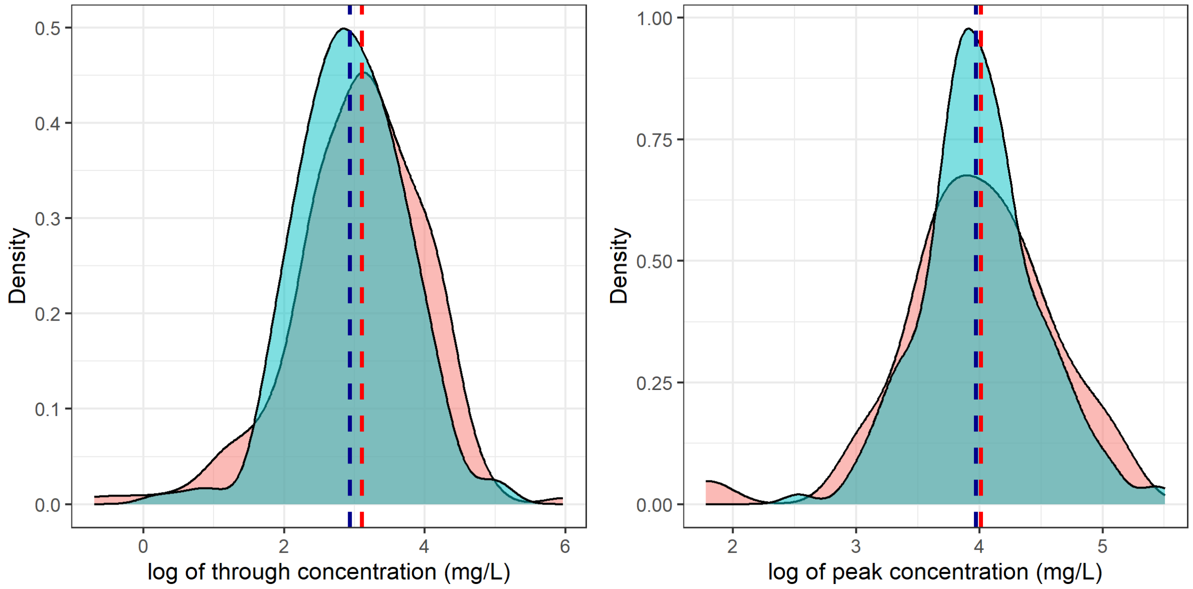


**Figure S1:** distribution of the natural logarithm of through concentrations (left, first measurements are censored) and peak concentrations (right). Dashed line: distribution median. Blue: MIPD group. Red: control group.

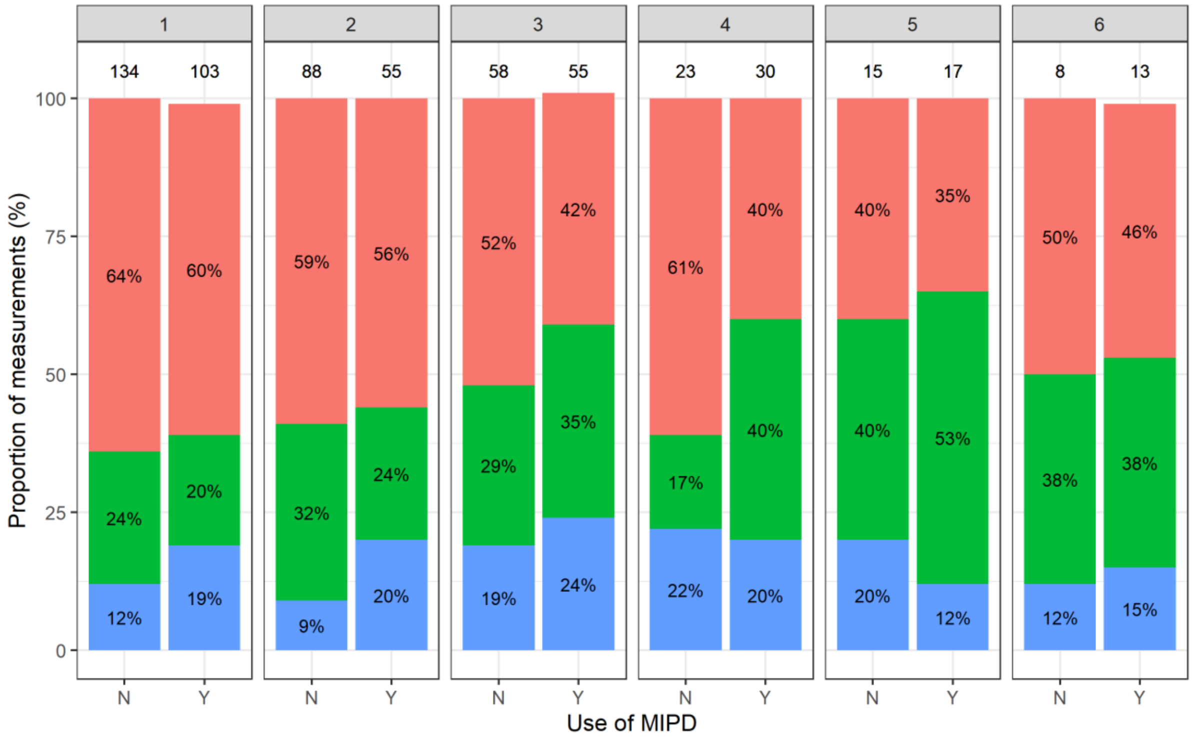


**Figure S2:** sensitivity analysis of proportion of measured through concentration in regard to the therapeutic window, stratified by group and TDM occasion. Through concentrations measured on blood samples collected more than 2 hours before the next administration were censored. Blue: through concentration under 10mg/L. Green: through concentration at target. Red: through concentration above 20mg/L. Absolute counts of patients of each group for each occasion are showed on top of the stacked bars. Percentages of each category of target attainment are showed inside the bars. Occasions later than 6 have been censored due to sparsity of data.

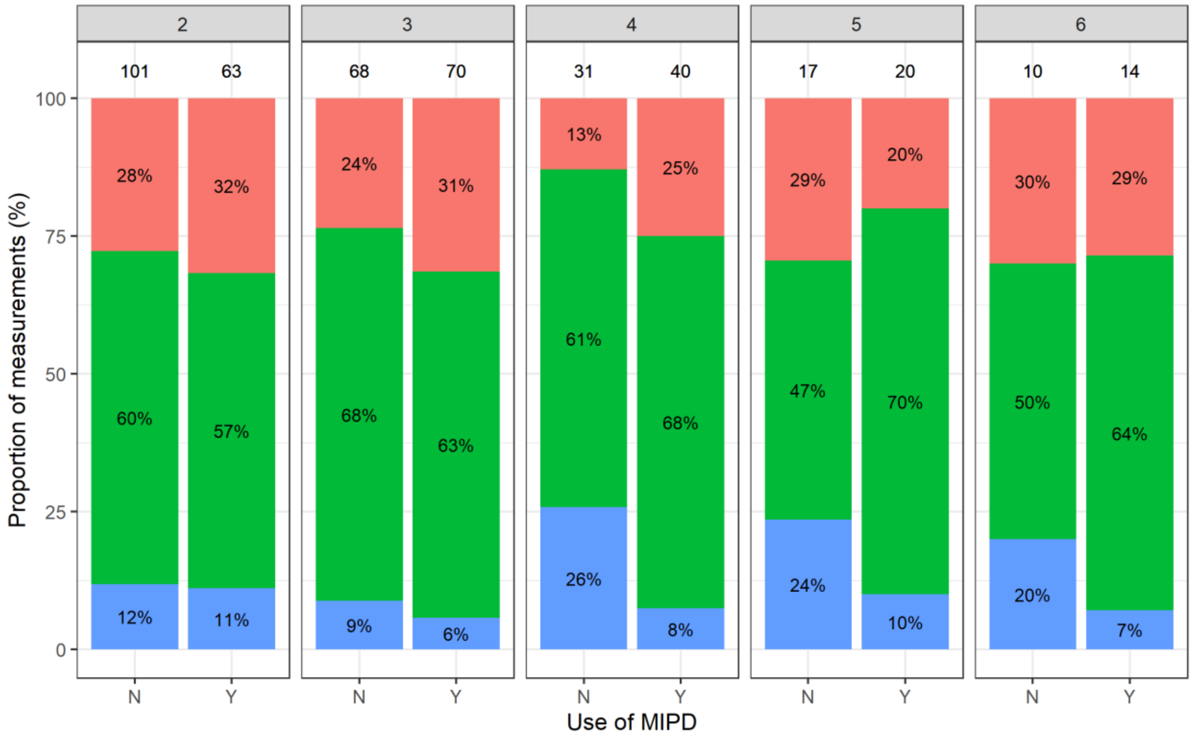


**Figure S3:** Proportions of dosage changes since previous observation. Blue: increased dosage. Green: unchanged dosage. Red: reduced dosage. Absolute counts of patients of each group for each occasion are showed on top of the stacked bars. Percentages of each category of target attainment are showed inside the bars. Occasions later than 6 have been censored due to sparsity of data.

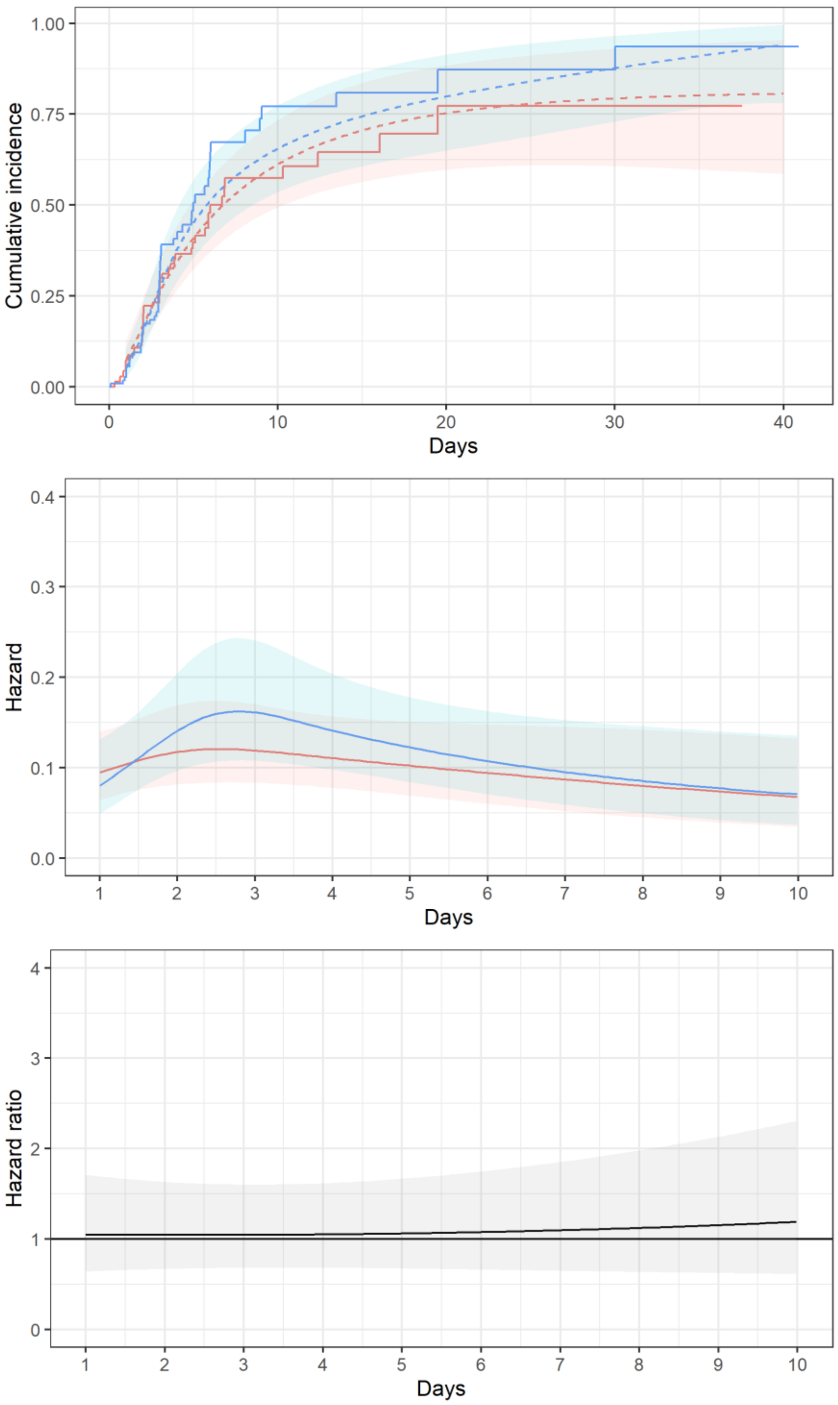


**Figure S4:** sensitivity analysis of modelling of time to target attainment. Residual concentrations measured on blood samples drew at least 2 hours before the real pharmacokinetic through point have been censored. Blue: MIPD group. Red: control group. **Top panel:** cumulative incidence of target attainment. Solid line: Kaplan-Meyer estimator. Dashed line: cumulative incidence predicted under the rate flexible model. Coloured area: 95% confidence interval of prediction. **Middle panel:** target attainment rate predicted by the flexible model. Coloured area: 95% confidence interval of prediction. **Bottom panel:** hazard ratio predicted under the rate flexible model. Grey area: 95% confidence interval of prediction. Prediction of hazard and hazard ratio later than day 10 are not shown, due to sparsity of input data.

### STROBE checklist

| | Item | Recommendation | Lines |
|---|---|---|---|
| **Introduction** | | | |
| Background/rationale | 2 | Explain the scientific background and rationale for the investigation being reported | 13 – 31 |
| Objectives | 3 | State specific objectives, including any prespecified hypotheses | 34 – 35 |
| **Methods** | | | |
| Study design | 4 | Present key elements of study design early in the paper | 38 |
| Setting | 5 | Describe the setting, locations, and relevant dates, including periods of recruitment, exposure, follow-up, and data collection | 44 – 45 |
| Participants | 6 | (*a*) Give the eligibility criteria, and the sources and methods of selection of participants. Describe methods of follow-up | 46 – 48 |
| | | (*b*) For matched studies, give matching criteria and number of exposed and unexposed | |
| Variables | 7 | Clearly define all outcomes, exposures, predictors, potential confounders, and effect modifiers. Give diagnostic criteria, if applicable | 164 – 168 |
| Data sources/ measurement | 8* | For each variable of interest, give sources of data and details of methods of assessment (measurement). Describe comparability of assessment methods if there is more than one group | 106 – 112 |
| Bias | 9 | Describe any efforts to address potential sources of bias | 174 – 190 |
| Study size | 10 | Explain how the study size was arrived at | 49 – 54 |
| Quantitative variables | 11 | Explain how quantitative variables were handled in the analyses. If applicable, describe which groupings were chosen and why | 142 – 143 |
| Statistical methods | 12 | (*a*) Describe all statistical methods, including those used to control for confounding | 145 – 163 |
| | | (*b*) Describe any methods used to examine subgroups and interactions | |
| | | (*c*) Explain how missing data were addressed | 114 – 116 |
| | | (*d*) If applicable, explain how loss to follow-up was addressed | |
| | | (*e*) Describe any sensitivity analyses | 174 – 190 |
| **Results** | | | |
| Participants | 13 | (a) Report numbers of individuals at each stage of study—eg numbers potentially eligible, examined for eligibility, confirmed eligible, included in the study, completing follow-up, and analysed | 196 – 203 |
| | | (b) Give reasons for non-participation at each stage | Figure 1 |
| | | (c) Consider use of a flow diagram | |
| Descriptive data | 14 | (a) Give characteristics of study participants (eg demographic, clinical, social) and information on exposures and potential confounders | 204 – 210 |
| | | (b) Indicate number of participants with missing data for each variable of interest | |
| | | (c) Summarise follow-up time (eg, average and total amount) | 205 – 206 |
| Outcome data | 15 | Report numbers of outcome events or summary measures over time | 225 – 230 |
| Main results | 16 | (*a*) Give unadjusted estimates and, if applicable, confounder-adjusted estimates and their precision (eg, 95% confidence interval). Make clear which confounders were adjusted for and why they were included | 243 – 250 |
| | | (*b*) Report category boundaries when continuous variables were categorized | |
| | | (*c*) If relevant, consider translating estimates of relative risk into absolute risk for a meaningful time period | |
| Other analyses | 17 | Report other analyses done—eg analyses of subgroups and interactions, and sensitivity analyses | 248 – 286 |
| **Discussion** | | | |
| Key results | 18 | Summarise key results with reference to study objectives | 297 – 302 |
| Limitations | 19 | Discuss limitations of the study, taking into account sources of potential bias or imprecision. Discuss both direction and magnitude of any potential bias | 361 – 392 |
| Interpretation | 20 | Give a cautious overall interpretation of results considering objectives, limitations, multiplicity of analyses, results from similar studies, and other relevant evidence | 307 – 316 |
| Generalisability | 21 | Discuss the generalisability (external validity) of the study results | 339 – 360 |
| **Other information** | | | |
| Funding | 22 | Give the source of funding and the role of the funders for the present study and, if applicable, for the original study on which the present article is based | 399 |
